# SibBMS: Siberian Brain Multiple Sclerosis Dataset with lesion segmentation and patient meta information

**DOI:** 10.1101/2025.08.14.25333741

**Authors:** Bair N. Tuchinov, Anna I. Prokaeva, Lyubov M Vasilkiv, Yuliya A. Stankevich, Denis S. Korobko, Nadezhda A. Malkova, Andrey A. Tulupov

## Abstract

Multiple sclerosis (MS) is a chronic inflammatory neurodegenerative disorder of the central nervous system (CNS) and represents the leading cause of non-traumatic disability among young adults. Magnetic resonance imaging (MRI) has revolutionized both the clinical management and scientific understanding of MS, serving as an indispensable paraclinical tool. Its high sensitivity and diagnostic accuracy enable early detection and timely therapeutic intervention, significantly impacting patient outcomes. Recent technological advancements have facilitated the integration of artificial intelligence (AI) algorithms for automated lesion identification, segmentation, and longitudinal monitoring. The ongoing refinement of deep learning (DL) and machine learning (ML) techniques, alongside their incorporation into clinical workflows, holds great promise for improving healthcare accessibility and quality in MS management. Despite the encouraging performance of DL models in MS lesion segmentation and disease progression tracking, their effectiveness is frequently constrained by the scarcity of large, diverse, and publicly available datasets. Open-source initiatives such as MSLesSeg, MS-Baghdad, MS-Shift, and MSSEG-2 have provided valuable contributions to the research community. Building upon these foundations, we introduce the SibBMS dataset to further advance data-driven research in MS. In this study, we present the SibBMS dataset, a carefully curated, open-source resource designed to support MS research utilizing structural brain MRI. The dataset comprises imaging data from 93 patients diagnosed with MS or radiologically isolated syndrome (RIS), alongside 100 healthy controls. All lesion annotations were manually delineated and rigorously reviewed by a three-tier panel of experienced neuroradiologists to ensure clinical relevance and segmentation accuracy. Additionally, the dataset includes comprehensive demographic metadata—such as age, sex, and disease duration—enabling robust stratified analyses and facilitating the development of more generalizable predictive models. Our dataset is available via a request-access form at https://forms.gle/VqTenJ4n8S8qvtxQA.

## 1. Introduction

Multiple sclerosis (MS) constitutes a chronic inflammatory neurodegenerative disorder of the central nervous system (CNS) that represents the primary cause of non-traumatic disability in young adults [6]. Hallmark pathological features include inflammation, demyelination, gliosis, and neuronal loss. Myelinated axons within the CNS serve as the principal targets of MS pathogenesis, resulting in heterogeneous damage to both myelin and axonal integrity. This process drives the formation of focal white matter lesions in the brain and spinal cord [4, 11].

Magnetic resonance imaging (MRI) has profoundly advanced both clinical management and scientific investigation of MS, serving as a cornerstone paraclinical tool. Its high sensitivity and diagnostic accuracy facilitate early detection and therapeutic intervention [12]. Precise identification of lesions within characteristic MS regions—including periventricular zones, cortical/juxtacortical areas, infratentorial structures, and the spinal cord — is critical for diagnostic confirmation [14]. Concurrently, longitudinal assessment of lesion dynamics is essential for guiding therapeutic strategies and enabling timely treatment modifications [13]. Recent technological advancements have enabled the integration of artificial intelligence (AI) algorithms for automated lesion identification, segmentation, and longitudinal comparison [1]. Continued refinement of deep learning (DL) and machine learning (ML) methodologies, coupled with their translation into clinical workflows, promises to enhance healthcare accessibility and quality. This study Name1 and Name2, presents a comprehensive characterization of the SibBMS database. This resource was developed to investigate the progression and evolution of morpho-functional alterations in brain tissue during demyelinating diseases while also enabling the training of AI algorithms for demyelination foci recognition. Subsequent application of this database is anticipated to accelerate the validation and clinical deployment of machine learning models.

## 2. Dataset Summary

This study encompasses both analytical and experimental (clinical) components. The analytical phase involved the systematic collection of information, including general and technical details pertinent to the acquisition of medical data—specifically, technical specifications and relevant radiological parameters of imaging equipment and protocols. Furthermore, a comprehensive review and comparative analysis of existing MS datasets and their respective data collection methodologies was conducted, with a focus on approaches utilized in MS diagnostic research. The experimental (clinical) component was dedicated to the evaluation and justification of the methodologies employed for data acquisition, preprocessing, annotation, and curation. This included the development and validation of annotation protocols, the assessment of annotator qualifications, and a detailed cohort analysis with explicit inclusion and exclusion criteria.

Key Features of the Collected Dataset

▪ Comprehensive MRI Protocols. The dataset comprises a full suite of core MRI sequences, including T2-FLAIR, T2-weighted imaging (T2-WI), T1-weighted imaging (T1-WI), and contrast-enhanced T1-weighted imaging (CE T1-WI). Notably, CE T1-WI facilitates the identification of active demyelinating lesions. Some example of visualizations on Fig 1.
▪ Extensive Meta- and Clinical Data. Each case is accompanied by essential demographic and clinical metadata, such as age, sex, date of diagnosis, age at disease onset, disease duration, disease subtype (relapsing-remitting MS [RRMS], secondary progressive MS [SPMS], primary progressive MS [PPMS], radiologically isolated syndrome (RIS), disease-modifying therapy (DMT) regimen, and Expanded Disability Status Scale (EDSS) score. This information is critical for the differential diagnosis of demyelinating disorders.
▪ Well-Characterized Control Group. The dataset includes an alternative control cohort of 100 healthy volunteers with no history of neurological disorders, normal neurological status, absence of cardiovascular and cerebrovascular diseases, and no MRI evidence of volumetric or focal brain lesions or cerebral hemodynamic abnormalities. MRI protocols for the control group include T2-FLAIR, T2, and T1 sequences. The inclusion of this control cohort is essential for the development and validation of intelligent diagnostic systems for MS.

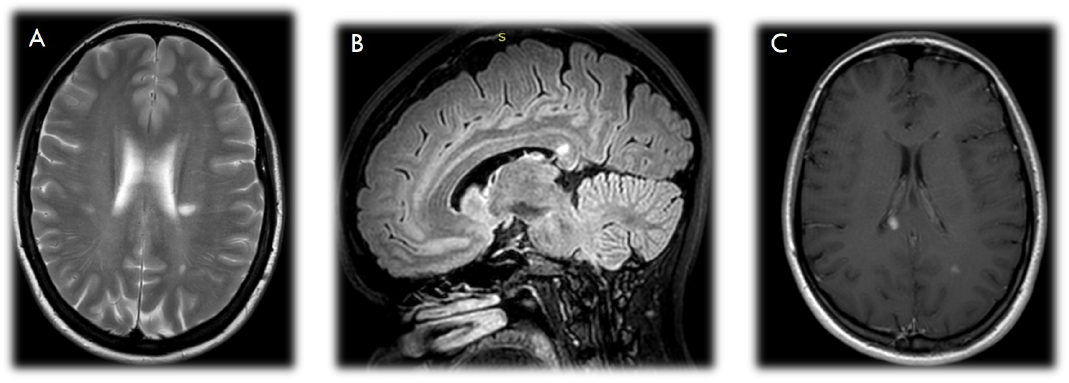

### 2.1 Dataset Description

The dataset compiled for this study comprises the following components:

▪ Preprocessed Imaging Data. The collection includes preprocessed MRI scans from 93 patients diagnosed with MS and 100 healthy control subjects. All imaging data have been converted from the original DICOM format to the standardized NIfTI format to facilitate interoperability and downstream analysis.
▪ Comprehensive Clinical Metadata. Accompanying the imaging data is a detailed table containing clinical and demographic parameters for each subject. This metadata encompasses variables such as age, sex, disease duration, disease subtype, therapeutic interventions, and relevant clinical scores.
▪ Annotated Lesion Segmentations. The dataset also provides a set of 10 manually annotated lesion segmentations, serving as reference examples for the development and validation of automated lesion detection and segmentation algorithms.

#### 2.1.1 Main demographic and clinical cohort’s characteristics

Data from patients diagnosed with MS or RIS, aged between 18 and 70 years, were retrospectively collected from two institutions: the Novosibirsk Regional Multiple Sclerosis and Other Autoimmune Diseases of the Nervous System Center (State Novosibirsk Regional Clinical Hospital) and the Institute of the International Tomography Center of the Siberian Branch of the Russian Academy of Sciences. The data collection period spanned from 2019 to 2024. All procedures adhered to international ethical standards for research involving human subjects, including obtaining informed consent from participants and ensuring confidentiality in accordance with the Declaration of Helsinki (World Medical Association, 2000 amendment) and the Russian Federation’s clinical practice regulations (Order of MELBA Journal Sample Article the Ministry of Health of Russia No. 266, dated 19 June 2003). The study protocol was approved and overseen by the Local Ethics Committee of the Institute of the International Tomography Center, SB RAS (protocol No.13, dated 11 June 2019).

#### 2.1.2 Inclusion and Exclusion Criteria

Patients were included if they met one of the following diagnostic criteria:

1. Diagnosis of MS according to the 2017 revised McDonald criteria,
2. Diagnosis of radiologically isolated syndrome (RIS) according to RIS criteria.

Additionally, patients were required to be aged 18 years or older.

Exclusion criteria comprised:

1. Contraindications to contrast-enhanced brain MRI, and
2. Presence of brain lesions attributable to etiologies other than MS (ischemic, traumatic etc.)

#### 2.1.3 Clinical Assessment

A comprehensive review of patients’ medical histories was conducted by neurologists specializing in MS. Neurological examinations were performed by certified raters to determine the Expanded Disability Status Scale (EDSS) score. The EDSS is a clinician-administered scale designed to quantify neurological impairment and disability in MS, assessing multiple functional systems of the central nervous system. The scale ranges from 0 (normal neurological function) to 10 (death due to MS), with increments of 0.5 from EDSS 1 onwards [5, 8].

#### 2.1.4 Cohort Composition and Data Quality Control

To ensure compliance with inclusion and exclusion criteria, data from over 300 patients were initially screened. After rigorous evaluation, the final dataset comprised 93 patients diagnosed with MS, or RIS. Detailed methodologies for the statistical analysis of demographic and clinical variables are provided in the Methods section. The demographic and clinical characteristics of the cohort are summarized in Table 1 and Table 2.

**Table 1:** Descriptive statistics for categorical variables

| Variables | Categories | Abs. | % | 95% CI |
| --- | --- | --- | --- | --- |
| Gender | female | 57 | 61.3 | 50.6 – 71.2 |
|  | male | 36 | 38.7 | 28.8 – 49.4 |
| Disease subtype | RRMS | 57 | 61.3 | 50.6 – 71.2 |
|  | SPMS | 24 | 25.8 | 17.3 – 35.9 |
|  | PPMS | 5 | 5.4 | 1.8 – 12.1 |
|  | RIS | 7 | 7.5 | 3.1 – 14.9 |
| DMT | not used | 20 | 21.5 | 13.7 – 31.2 |
|  | used | 73 | 78.5 | 68.8 – 86.3 |

**Table 2:** Descriptive statistics for quantitative variables

| Variables | M ± SD / Me | 95% CI / Q<sub>1</sub>–Q<sub>3</sub> | n | min | max |
| --- | --- | --- | --- | --- | --- |
| Age (years) | 40.9 ± 11.5 | 38.5 – 43.3 | 93 | 19.0 | 71.0 |
| Age of onset (years) | 31.7 ± 11.3 | 29.3 – 34.0 | 93 | 9.2 | 65.2 |
| Disease duration (years) | 8.00 | 5.00 – 13.00 | 93 | 0.00 | 46.00 |
| EDSS (scores) | 2.0 | 1.5 – 3.0 | 93 | 0.0 | 7.5 |

#### 2.1.5 Data Acquisition Details & Equipment Specifications

The study was performed using two high-field magnetic resonance imaging (MRI) systems manufactured by Philips: the Achieva 1.5 Tesla (T) scanner and the Ingenia 3 Tesla scanner. To exclude or confirm brain pathology, each imaging session commenced with a standardized MRI protocol comprising acquisition of T1-weighted (T1-WI) and T2-weighted (T2-WI) sequences, along with fluid-attenuated inversion recovery (FLAIR) images for cerebrospinal fluid (CSF) signal suppression. (Table 3). Measurements were carried out at the Center of Collective Use “Mass spectrometric investigations” SB RAS”.

**Table 3:** Characteristics of the used magnetic resonance imaging protocols for brain examination

MRI scanner: “Achieva Philips” 1.5 Tesla
|  | T1WI_TSE | T2WI_TSE | T2WI_FLAIR | CE_T1WI_TSE |
| --- | --- | --- | --- | --- |
| Slice orientation | Sagittal | Axial | Frontal | Sagittal |
| Pulse sequence | TSE | TSE | TSE_IR | TSE |
| TR/TE | 549 / 15 | 5148 / 100 | 11675 / 140TI 2800 ms | 549 / 15 |
| Matrix | 256 × 180 | 372 × 247 | 300 × 165 | 256 × 180 |
| Voxel spacing (mm) | 0.94 × 0.94 × 5.0 | 0.49 × 0.49 × 4.0 | 0.84 × 0.84 × 4.0 | 0.94 × 0.94 × 5.0 |
| Slice thickness | 5 | 4 | 4 | 5 |
| NSA | 2 | 2 | 2 | 2 |
| Scan time | 1 min 11 s | 2 min 29 s | 2 min 55 s | 1 min 11 s |

MRI scanner: Ingenia Philips 3 Tesla
|  | 3D_T1WI_TFE | T2WI_TSE | 3D_T2WI_FLAIR | CE_3D_T1WI_TFE |
| --- | --- | --- | --- | --- |
| Slice orientation | Axial | Axial | Axial | Axial |
| Pulse sequence | TFE | TSE | TSE | TFE |
| TR/TE | 7.9 / 3.9 | 3000 / 80 | 4800 / 331TI 1650 | 7.9 / 3.9 |
| Matrix | 252 × 227 | 420 × 272 | 204 × 196 | 252 × 227 |
| Voxel spacing (mm) | 0.87 × 0.87 × 1.0 | 0.4 × 0.4 × 4.0 | 0.72 × 0.72 × 0.6 | 0.87 × 0.87 × 1.0 |
| Slice thickness | 1.2 | 4 | 1.2 | 1.2 |
| NSA | 2 | 2 | 2 | 2 |
| Scan time | 3 min 1 s | 3 min 36 s | 6 min 10 s | 3 min 1 s |

The imaging protocol encompassed comprehensive coverage of the entire brain, including the frontal, parietal, occipital, and temporal lobes; basal ganglia and internal capsules; corpus callosum; cerebral peduncles; midbrain; pons; cerebellar peduncles and hemispheres; medulla oblongata; and the initial segments of the cervical spinal cord. Both patients with demyelinating diseases and healthy control volunteers underwent this standard imaging protocol, supplemented by additional sequences as required, within a single session. The total duration of each MRI examination, inclusive of all routine sequences, was approximately 25 minutes per subject. The State Novosibirsk Regional Clinical Hospital and the Institute of the International Tomography Center authorized the release of the data under a Creative Commons Attribution 3.0 International license (CC BY-NC-SA 3.0).

### 2.2 Annotation and Segmentation

The image analysis workflow for each subject involved the following sequential steps:

1. Qualitative Slice-by-Slice Assessment Pre- and post-contrast T1-WI, T2-WI, and FLAIR images were systematically reviewed on a slice-by-slice basis to evaluate focal brain lesions indicative of demyelination.
2. Data Conversion and Image Registration Original DICOM files were converted to the NIfTI format to facilitate advanced image processing. The datasets were imported into 3D Slicer software (version 5.4.0). All images were spatially registered with a reference FLAIR volume (NMRI225 Flair.nii), generating a registered dataset denoted as “N reg flair,” where “N” corresponds to the patient identifier. Subsequently, T2-WI and both pre- and post-contrast T1-WI sequences were co-registered to the “N reg flair” volume to ensure spatial congruence across modalities.
3. Lesion Segmentation and Annotation Segmentation focused on delineating demyelinating lesions categorized by anatomical location: subcortical, juxtacortical, periventricular, and infratentorial foci. FLAIR images served as the primary modality for lesion identification and segmentation, while T1-WI and T2-WI sequences were utilized to refine lesion boundaries and support lesion scoring. The example of annotation on Fig 2. Manual segmentation and annotation were conducted and validated through a rigorous multi-tiered expert review process: **First Tier** Initial segmentation was performed by a radiologist with three years of experience, who conducted a comprehensive slice-by-slice examination for all subjects. **Second Tier** Two senior radiologists, each with over ten years of experience, reviewed and corrected the initial segmentations to address inaccuracies or omissions. **Third Tier** A neurologist with extensive clinical expertise in demyelinating diseases performed a final verification and correction of the segmentations to ensure clinical validity and diagnostic accuracy. This three-level annotation and verification protocol was designed to maximize segmentation precision and reliability, integrating imaging findings with relevant clinical and demographic metadata.
4. Segmentation Export Finalized lesion segmentations were extracted from the native imaging series and saved in the NIfTI format for subsequent quantitative analysis and algorithm training.

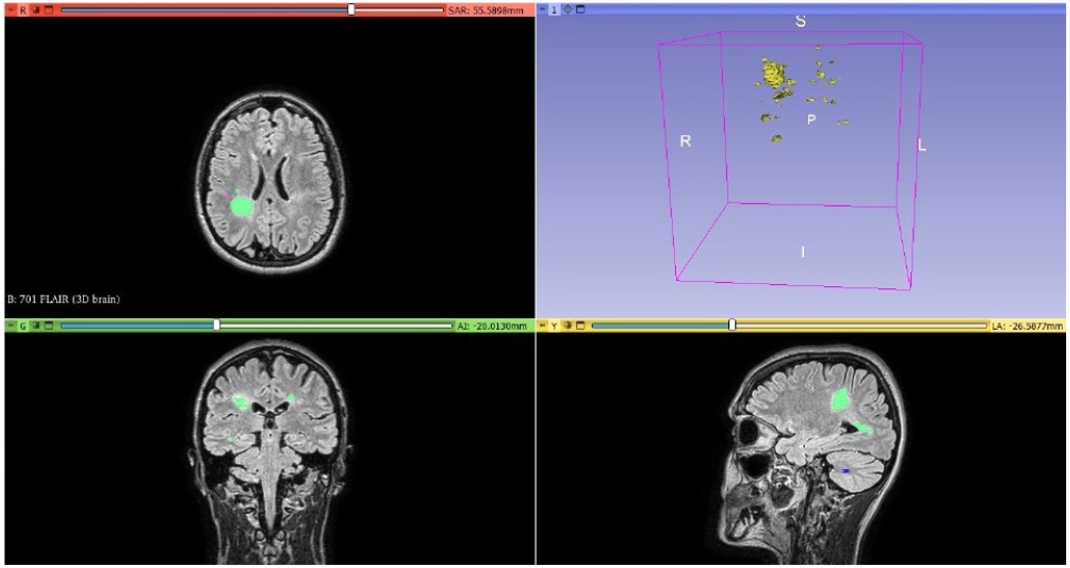

## 3. Methods

Data collection, preprocessing, annotation, and storage were facilitated using proprietary tools developed by our team: the AI Service for Preprocessing and Diagnostics of MRI Images and the Kappa Framework for data curation and management [10]. The AI Service for Preprocessing and Diagnostics of MRI Images implements a standardized pipeline comprising the following steps:

▪ Standardization of Input Data Structure: Ensuring uniform formatting and organization of incoming MRI data to enable consistent downstream processing.
▪ DICOM Compliance Verification and Metadata Extraction: Validating that all imaging data conform to the DICOM standard and extracting relevant metadata critical for analysis.
▪ Conversion from DICOM to NIfTI Format and Data Validation: Transforming imaging data into the NIfTI format, followed by rigorous validation to ensure data integrity and completeness.
▪ Technical Quality Assessment: Automated evaluation of image quality metrics to identify artifacts or inconsistencies that may affect analysis.
▪ Image Preprocessing: Application of preprocessing techniques such as normalization, registration, and noise reduction to prepare images for subsequent analysis. The Kappa Framework addresses the challenges of annotation curation, storage, and dataset utilization for AI model training and inference in clinical contexts. Its primary functionalities include:

1. Integration with Annotation and Machine Learning Tools: Seamless interoperability with labeling platforms such as CVAT and machine learning frameworks including TensorFlow, PyTorch, and OpenVINO.
2. Expert and Automated Labeling: While automated labeling methods have advanced, manual expert annotation remains essential. The framework incorporates structured workflows to mitigate inter-annotator variability and ensure labeling consistency.
3. Cross-Labeling by Multiple Experts: Facilitates independent annotation of the same dataset by multiple experts or models, a critical process in domains demanding high accuracy and specialized expertise, such as medical imaging diagnostics and legal document classification.
4. Consensus Labeling: Implements protocols to reconcile divergent annotations from multiple experts, producing a unified ground truth label for each data point.
5. Quality Control of Annotations and Datasets: Conducts systematic reviews to detect and resolve annotation discrepancies, thereby maintaining dataset fidelity.
6. Secure Data Storage: Provides compliant, secure storage solutions to protect sensitive clinical and imaging data.

### 3.1 Statistical Analysis

Statistical analyses of patient demographic and clinical data were performed using StatTech v. 4.8.5 (StatTech LLC, Russia). The distribution of quantitative variables was assessed for normality using the Kolmogorov-Smirnov test. Variables conforming to a normal distribution are reported as mean (M) ± standard deviation (SD), with 95% confidence intervals (95% CI) provided to indicate estimated precision. Non-normally distributed quantitative variables are summarized using median (Me) and interquartile range (Q1–Q3). Categorical variables are described using absolute counts and relative frequencies, with 95% confidence intervals for proportions calculated via the Clopper-Pearson exact method. Using these methods, a descriptive statistical analysis of the study cohort was performed. The demographic and clinical characteristics of the cohort are summarized in Table 1 and Table 2 (see above).

### 3.2 Overview of Open-Source Multiple Sclerosis MRI Datasets

The MS-Shift[7] and MSSEG-2 [2] datasets include the MRI image information and segmentation label masks. On MSLesSeg and MS-Baghdad added some patient metadata and clinical information related to disease status.

The Baghdad dataset [9] comprises two-dimensional FLAIR MRI scans from 60 patients diagnosed with multiple sclerosis (MS), collected at the MS Clinic of Baghdad Teaching Hospital, Medical City Complex, Baghdad, Iraq. The cohort includes 46 females and 14 males, with a mean age of 33 years (range: 15–56 years). MRI acquisitions were performed between 2019 and 2020 using 1.5 Tesla scanners across 20 centers. The mean Expanded Disability Status Scale (EDSS) score was 2.3, ranging from 0 to 6; notably, 78% of patients exhibited an EDSS score below 4, while the remaining 22% had scores of 4 or greater. All patients received a confirmed diagnosis of MS by neurologists at the MS Clinic. Comprehensive patient metadata accompany the imaging data, including demographic information (age, gender), comorbidities, age at disease onset, presenting symptoms, and medication regimens. Clinical data further encompass detailed neurological examination results, assessing functional domains such as pyramidal and cerebellar systems, brainstem function, sensory modalities, sphincter control, visual function, mental status, speech, motor coordination, gait, bowel and bladder function, optic disc evaluation, visual fields, nystagmus, eye movements, swallowing, and EDSS scores.

The MSLesSeg dataset [3] includes MRI data from 75 patients aged 18 to 59 years, with a mean baseline age of 37 years (±10.3). The cohort consists of 48 females and 27 males. MRI scans were retrospectively collected at multiple time points per patient, ranging from one to four observations: 50 patients had a single observation, 15 had two, 5 had three, and 5 had four, totaling 115 imaging series. Each observation includes three imaging sequences: T1-weighted (T1-w), T2-weighted (T2-w), and FLAIR. The dataset is partitioned into training and testing subsets. The training set comprises 53 patients with one to four observations, predominantly relapsing-remitting multiple sclerosis (RRMS; n=50) and a minority with secondary progressive multiple sclerosis (SPMS; n=3). The test set includes 22 patients, each with a single observation, primarily RRMS (n=21) and one case of primary progressive multiple sclerosis (PPMS). Supplementary materials provide detailed patient-level data, including MS subtype, age, EDSS scores, lesion volume, and lesion count.

## 4. Conclusions

A comprehensive database of MRI images from patients with diverse forms of MS has been established, incorporating extensive patient metadata and clinical information related to disease status. Expert manual segmentation was performed on preprocessed 3D FLAIR images using the 3D Slicer software package (version 5.4.0). Radiologists systematically imported the imaging studies into 3D Slicer, conducted preliminary quality assessments, and optimized image contrast. Initial registration aligned FLAIR images to a reference volume (NMRI225 Flair), followed by subsequent registration of T2-weighted and pre- and post-contrast T1-weighted sequences relative to the registered FLAIR images. Lesion labeling adhered to the McDonald criteria, categorizing lesions by anatomical location into juxtacortical and subcortical, periventricular, and infratentorial groups. Radiologists manually delineated lesions on each slice using dedicated segmentation tools within 3D Slicer. This multi-step annotation process ensured precise and consistent lesion identification. The resulting dataset and annotations provide a valuable resource for the development and validation of artificial intelligence and computer vision algorithms aimed at the dynamic assessment of radiological and clinical manifestations of MS. These tools have the potential to enhance diagnostic accuracy, monitor disease progression, and support personalized therapeutic strategies.

## Acknowledgments

This work was supported by a grant from the Russian Science Foundation (RSF 23-15-00377)

## Ethical Standards

The work follows appropriate ethical standards in conducting research and writing the manuscript, following all applicable laws and regulations regarding treatment of human subjects in accordance with the Declaration of Helsinki (World Medical Association, 2000 amendment) and the Russian Federation’s clinical practice regulations (Order of the Ministry of Health of Russia No. 266, dated 19 June 2003), with the protocol by the Local Ethics Committee of the Institute of the International Tomography Center, SB RAS (protocol No.13, dated 11 June 2019).

## Conflicts of Interest

The conflicts of interest have not been entered yet.

## Data availability

The SibBMS dataset is available via a request-access form at https://forms.gle/VqTenJ4n8S8qvtxQA.

## Notes

### Competing Interest Statement

The authors have declared no competing interest.

### Author Declarations

The study protocol was approved and overseen by the Local Ethics Committee of the Institute of the International Tomography Center, SB RAS (protocol No.13, dated 11 June 2019).

